# Effect of *MAOA* DNA methylation on human *in vivo* protein expression measured by [^11^C]harmine PET in healthy and depressed individuals

**DOI:** 10.1101/2022.03.29.22273110

**Authors:** Patricia A. Handschuh, Matej Murgaš, Chrysoula Vraka, Lukas Nics, Annette M. Hartmann, Edda Winkler-Pjrek, Pia Baldinger-Melich, Wolfgang Wadsak, Dietmar Winkler, Marcus Hacker, Dan Rujescu, Katharina Domschke, Rupert Lanzenberger, Marie Spies

## Abstract

Epigenetic modifications, such as DNA methylation, are understood as an intermediary between environmental factors affecting disease risk and pathophysiologic changes to brain structure and function. Cerebral monoamine oxidase A (MAO-A) levels are altered in depression, as are DNA methylation levels within the *MAOA* gene, particularly in the promoter / exon I / intron I region. An effect of *MAOA* methylation on peripheral protein expression was shown, but the extent to which methylation affects brain MAO-A levels is not fully understood. Here, the influence of average and CpG site-specific *MAOA* promoter / exon I / intron I region DNA methylation on global MAO-A distribution volume (V_T_), an index of MAO-A density, was assessed via [^11^C]harmine positron emission tomography in 22 patients suffering from winter-type seasonal affective disorder and 30 healthy controls. No significant influence of *MAOA* DNA methylation on global MAO-A V_T_ was found, despite correction for health status (patients vs. controls), sex, season (methylation analysis in spring / summer vs. fall / winter) and *MAOA* variable number of tandem repeat genotype (VNTR; high vs. low expression groups). However, in female subjects, season affected average DNA methylation, with higher levels in spring and summer (p_uncorr_ = 0.03). We thus did not find evidence for an effect of *MAOA* DNA methylation on brain MAO-A V_T_. In contrast to a previous study that demonstrated an effect of the methylation of a *MAOA* promoter region located further 5’ on brain MAO-A, in the present study *MAOA* methylation appears to affect brain protein levels to a limited extent. The observed effect of season on methylation levels is in accordance with extensive evidence for seasonal effects within the serotonergic system.

**Clinicaltrials.gov Identifier:** NCT02582398

**EUDAMED Number:** CIV-AT-13-01-009583

## 1 Introduction

As the enzyme primarily responsible for degradation of serotonin, but also dopamine and norepinephrine, monoamine oxidase A (MAO-A) is integral to monoaminergic homeostasis in the human brain. Alterations to MAO-A function have been associated with risk (1, 2), pathophysiology and treatment (3) of a range of psychiatric illnesses. These include affective (4), anxiety (5), obsessive compulsive (OCD) (6), substance use (7) and personality disorders (8). In depression, MAO-A hyperactivity is thought to result in reduced serotonin signaling. This phenomenon is a key component of the serotonergic hypothesis of depression (4). Information provided by peripheral assessments of MAO function, for example via platelets, is limited, as platelet MAO expression is restricted to that of MAO-B, while the brain expresses both isoenzymes (9). Positron emission tomography (PET) with [^11^C]harmine provides specific *in vivo* information on brain MAO-A density and distribution (10, 11). In particular, [^11^C]harmine PET studies have provided evidence for changes to MAO-A total distribution volume (V_T_), an index of protein levels, in major depression (4).

Changes to DNA methylation within the *MAOA* gene have been observed in affective disorders (12), anxiety (13, 14), OCD (15) and post-traumatic stress disorder (16), among others (17). *MAOA* DNA methylation is mediated both by risk factors for (18, 19) – as well as treatment of (14, 20) – various psychiatric disorders, suggestive of its role as an intermediate between environment and neurobiology (21). *In vitro* studies demonstrate a negative association between peripheral blood *MAOA* promoter / intron I / exon I methylation and protein function (14, 22). In a PET study utilizing *in vivo* [^11^C]clorgyline, a negative association between *MAOA* promoter methylation and brain MAO levels was demonstrated in healthy individuals (23). In theory, methylation of the promoter / exon I / intron I regions may be associated with particularly strong downregulation of transcription (21). *MAOA* promoter / exon I / intron I methylation also carries clinical significance and has been associated with a variety of psychiatric disorders (19, 24, 25). However, the effect of promoter / exon I / intron I methylation on human *in vivo* brain MAO-A levels in patients with depression, as measured by [^11^C]harmine, has yet to be assessed.

Seasonal affective disorder (SAD) is characterized by depressive symptoms in fall and winter and remission in spring and summer. Though evidence points towards a serotonergic pathophysiology (26, 27) and efficacy of MAO inhibitors such as moclobemide would point towards a role for MAO-A (28), a study by our group did not find changes to MAO-A V_T_ (29). In general, the role of epigenetic mechanisms in driving intra- and interpersonal differences in cerebral MAO-A levels is insufficiently understood.

Here we assess 1) the effect of average and CpG-specific *MAOA* promoter / exon I / intron I region DNA methylation on brain MAO-A V_T_ assessed with [^11^C]harmine PET in 30 healthy individuals and 22 patients with winter-type SAD. 2) We additionally take the seasonal pathophysiology of SAD into consideration by probing the impact of season on *MAOA* DNA methylation.

## 2 Materials and methods

### 2.1 Study design

The current study utilizes [^11^C]harmine PET and *MAOA* DNA methylation data from 30 healthy controls (HC) and 22 patients with winter type SAD (n = 52). Data was gleaned from a previously published randomized, longitudinal, double-blind study assessing changes to MAO-A V_T_ in SAD after treatment with bright light therapy (BLT) and across the seasons (29). This larger study comprised a screening visit, three PET measurements (PET1 - before treatment in fall / winter, PET2 - after treatment in fall / winter, and PET3 - in spring / summer) and a structural magnetic resonance imaging (MRI) scan. Study enrollment was completed by a follow-up visit. A blood draw for methylation analysis was performed either at the baseline PET measurement (PET1) or the follow-up visit. Here, we utilize the PET scan (PET1 - before treatment in fall / winter or PET3 - in spring / summer) that was performed closest to the time of methylation analysis for each subject (mean difference 27.63 ± SD 51.74 days between PET and blood draw). At the time of PET in spring / summer (PET3), patients were remitted from SAD and all subjects (i.e., patients and controls) had received treatment with BLT or placebo. The study was conducted in accordance with the Declaration of Helsinki, including all current revisions and the good scientific practice guidelines of the Medical University of Vienna. The protocol was approved by the ethics committee of the Medical University of Vienna (EK Nr.: 1681/2016) and registered at clinicaltrials.gov (NCT02582398).

### 2.2 Subjects

SAD patients were recruited via the respective outpatient clinic at the Department of Psychiatry and Psychotherapy, Medical University of Vienna. HC were recruited via advertisements in local newspapers, electronic media and through dedicated message boards at the General Hospital of Vienna and the Medical University of Vienna. The Structured Clinical Interview for DSM-IV Axis I disorders (SCID-I for DSM-IV) was used to diagnose unipolar, winter-type SAD and exclude psychiatric comorbidities in patients as well as to confirm psychiatric health in HC. In addition, to confirm (SAD patients) or exclude (HC) the diagnosis of SAD, all individuals completed the Seasonal Pattern Assessment Questionnaire (SPAQ) (30). Subjects were free from psychopharmacologic medication for the period of study participation and within 6 months prior to study enrollment. Severe somatic illness, neurologic comorbidities, current drug abuse, current smoking and pregnancy (female subjects) were excluded based on medical history, routine laboratory parameters (blood draw and urine tests), electrocardiography and physical examination performed at the screening visit. All individuals provided written informed consent and received financial reimbursement for their study participation.

### 2.3 Positron emission tomography

PET scans were performed with a GE Advance full-ring PET scanner (GE Medical Systems, Wukesha, WI) at the PET Center of the Department of Biomedical Imaging and Image-guided Therapy, Medical University of Vienna. [^11^C]harmine (7-[^11^C]methoxy-1-methyl-9H-[3,4-b]indole) synthesis and quality control were performed in line with the workflow presented by Philippe et al. (31). In a first step, a 5 min transmission scan was performed with ^68^GE rod sources for tissue attenuation. Dynamic PET scans started simultaneously with the intravenous bolus application of [^11^C]harmine (4.6 MBq/kg body weight) (4, 10, 11, 32, 33). All scans were acquired in 3D mode, collecting 51 successive time frames (12 × 5 sec, 6 × 10 sec, 3 × 20 sec, 6 × 30 sec, 9 × 1 min and 15 frames x 5 min), resulting in a total acquisition time of 90 min. Scans were reconstructed into 35 transaxial section volumes (128 × 128 matrix) utilizing an iterative filtered back-projection algorithm (FORE-ITER) with a spatial resolution of 4.36 mm full width at half maximum 1 cm next to the center of the field of view. Additionally, arterial blood samples for [^11^C]harmine quantification were drawn via automated blood sampling for the first 10 min of measurement (ALLOGG, Mariefred, Sweden), complemented by manual blood samples at standardized time points (at 5, 10, 20, 30, 45, 60 and 80 min after tracer application).

### 2.4 Magnetic resonance imaging

T1-weighted MR images (magnetization prepared rapid gradient echo (MPRAGE) sequence, 256 × 240 matrix, 1 × 1 mm voxel size, slice thickness 1.1 mm, 200 slices) were acquired using a 3 Tesla PRISMA MR Scanner (Siemens Medical, Erlangen, Germany) at the Medical University of Vienna.

### 2.5 MAO-A V_T_ quantification

Prior to quantification, each PET scan was spatially normalized to Montreal Neurological Institute (MNI) space using SPM12 (Wellcome Trust Centre for Neuroimaging, London, United Kingdom; http://www.fil.ion.ucl.ac.uk/spm/). In short, each PET was corrected for head motion and co-registered to the T1 structural image. Afterwards, each MR scan was normalized to MNI space utilizing a tissue probability map, producing the transformation matrix that was used to normalize the co-registered PET scan to MNI space.

Manually drawn arterial blood samples were processed according to the protocol published by Ginovart et al. (10). A gamma counter was cross-calibrated with the PET scanner as well as the automated arterial blood sampling system. To acquire non-metabolized [^11^C]harmine in arterial blood as a function of time, the arterial input function (AIF) was calculated as the product of whole blood activity (fit with 3 exponentials), plasma-to-whole blood ratio (linear fit), and the fraction of non-metabolized tracer concentration in arterial plasma (fit with Watabe function).

Logan plot was used for voxel-wise quantification of MAO-A V_T_ (31). Thereby, the estimated AIF and the time activity curve of thalamus, representing the high uptake region, were used. Regional V_T_ were extracted for frontal and temporal pole, anterior and posterior cingulate gyrus, thalamus, caudate, putamen, hippocampus and midbrain adopted from the Harvard-Oxford cortical structural atlas (http://fsl.fmrib.ox.ac.uk/fsl/fslwiki/Atlases) as well as striatum taken from an in-house atlas (32). Afterwards, a global region of interest (ROI) representing the weighted average of regional V_T_ was utilized, as relevant regional differences in methylation effects on MAO-A V_T_ were not hypothesized and because MAO-A V_T_ was highly correlated between regions (average correlation = 0.92 ± 0.04). PMOD 3.509 (PMOD Technologies Ltd., Zurich, Switzerland; www.pmod.com) was used to fit the arterial input function and for [^11^C]harmine quantification.

### 2.6 DNA sampling and isolation

Venous blood (approximately 24 ml from a cubital vein) was collected in EDTA blood tubes during the PET measurement or the follow-up examination and stored at −80 °C. DNA extraction was performed using the QIAamp DNA Blood Midi and Maxi Kit (Qiagen, Hilden, Germany) according to the manufacturer’s recommendations at the Department of Psychiatry, Psychotherapy and Psychosomatics of the University of Halle, Germany. Afterwards, DNA samples were again stored at −80 °C.

### 2.7 *MAOA* VNTR genotyping and methylation analysis

Subjects were genotyped for the *MAOA* variable number of tandem repeat (VNTR) promoter polymorphism containing 2, 3, 3.5, 4 or 5 copies of the repeated sequence (30), as this variant was shown *in vitro* to affect MAO-A expression (31–33). *MAOA* VNTR genotyping was performed at the Department of Psychiatry, Psychotherapy and Psychosomatics of the University of Halle, Germany. Briefly, 25μl PCR reactions containing 50 ng DNA, 10 pMol each of forward (5’-TGCTCCAGAAACATGAGCAC-3’) and reverse primers (5’-ATTGGGGAGTGTATGCTGGA-3’), 1U Taq polymerase, 10 mMol dNTPs, 15 mM ammonium sulfate, 60 mM Tris-HCl (pH 9.5), 1.5 mMol/μl MgCl2 were amplified in 35 cycles (94°C for 30s; 56°C for 30s; 72°C for 1min) after an initial denaturation step at 94 °C for 5 min and PCR fragments were resolved on a 2.5% agarose gel.

DNA methylation was then assessed at the Department of Psychiatry and Psychotherapy, University of Freiburg, Faculty of Medicine, Germany, via direct sequencing of bisulfite-converted DNA. Degree of methylation at 13 CpG sites located in an amplicon comprising promoter / exon I / intron I of *MAOA* (chromosome X, GRCh38.p2 Primary Assembly, NCBI Reference Sequence: NC_000023.11, 43656260–43656613) were analyzed individually. These CpG sites were numbered in accordance with prior studies on *MAOA* methylation in psychiatric diseases: CpG 1 = 43 656 316; CpG 2 = 43 656 327; CpG 3 = 43 656 362; CpG 4 = 43 656 368; CpG 5 = 43 656 370; CpG 6 = 43 656 383; CpG 7 = 43 656 386; CpG 8 = 43 656 392; CpG 9 = 43 656 398; CpG 10 = 43 656 427; CpG 11 = 43 656 432; CpG 12 = 43 656 514; CpG 13 = 43 656 553 (16, 34). For detailed information on *MAOA* methylation analysis, see Ziegler et al. (20). In addition, average methylation of these 13 CpG sites was determined.

### 2.8 Statistical analysis

Statistical tests were performed using SPSS version 28 for Windows (SPSS Inc., Chicago, Illinois; www.spss.com).

#### Analysis of *MAOA* DNA methylation

*MAOA* methylation data was available in 29 patients suffering from unipolar winter type SAD (mean age 32,83 ± SD 9.55 years, 18 females) and 44 healthy controls (mean age 34.27, ± SD 9.86 years, 26 females). Average *MAOA* methylation was defined as the average methylation of all pre-defined CpG sites (CpG 1-13).

Based on Shapiro–Wilk tests for normality and visual inspection, non-parametric testing was used for analysis of methylation data. Average *MAOA* methylation was used as the primary outcome parameter. Mann-Whitney U test was used to probe for differences in average *MAOA* methylation between males vs. females, SAD patients vs. healthy controls, the effect of season (methylation analysis in spring / summer, covering the period from April to September vs. autumn / winter, covering the period between October and March), and of VNTR high (3.5 and 4 repeats) vs. low expressing group (2, 3, and 5 repeats) (30). Inter-correlation of CpG sites ranged from −0.22 to 0.86 in females and from −0.24 to 0.77 in males (see supplement). Thus, exploratory analyses of individual CpG site methylation were performed utilizing the same steps as for average methylation.

#### Effect of *MAOA* DNA methylation on MAO-A V_T_

PET data was available in 22 SAD patients (mean age 33 ± 10.20 years, 14 females) and 30 HC (mean age 33.80 ± 9.76 years, 17 females). A general linear model (GLM) was utilized using global MAO-A V_T_ as the dependent variable. Sex, VNTR expression group, season and health status were used as fixed factors and age (Z-scored) as covariate to probe for their potential effect on MAO-A V_T_ for the subsequent analysis.

The effect of average *MAOA* methylation on global MAO-A V_T_ was assessed using GLM with global MAO-A V_T_ as the dependent variable and average methylation (Z-scored) as the covariate. Based on previous evidence for an effect of sex, season and depression on brain MAO-A V_T_ (29, 35, 36) a second GLM was computed additionally including sex, health status and season as fixed factors. In an exploratory manner, GLM analysis was repeated to assess individual CpG cites. Multiple testing was corrected using the Holm-Bonferroni method and significance was set at p < 0.05.

## 3 Results

### 3.1 Analysis of *MAOA* DNA methylation

Mann-Whitney U-test revealed a significant effect of sex on average methylation with higher levels in females (p_corr_ < 0.05), which was expected based on the X-chromosomal location of *MAOA*, X-chromosome hemizygosity in male, and X-chromosome inactivation in female cells. Hereafter, methylation data of male and female participants were investigated separately. No effect of health status or VNTR genotype on average methylation was detected. As shown in *Figure 1*, an effect of season on average methylation was observed in female subjects, showing higher methylation levels in women whose blood was drawn during spring or summer vs. autumn or winter (two-sided Mann-Whitney U-test, p_uncorr_ = 0.03). This finding did not survive correction for multiple comparisons. In exploratory analyses of individual CpGs, higher methylation at CpG 1 (p_uncorr_ = 0.004) and at CpG 3 (p_uncorr_ = 0.002) in healthy females compared to females with SAD was observed. In males, those suffering from SAD showed higher methylation at CpG 5 (p_uncorr_ = 0.025) than healthy controls. Additionally, significantly higher methylation was found at CpGs 2 (p_uncorr_ = 0.001), 3 (p_uncorr_ = 0.039), 5 (p_uncorr_ = 0.049), 7 (p_uncorr_ = 0.040) and 9 (p_uncorr_ = 0.023) in samples of females that were collected during spring or summer compared to those collected in autumn or winter. See *Table 1 and 2* for mean methylation levels.

**Table 1:** Mean *MAOA* promoter / exon I / intron I region DNA methylation levels (%) in females, average and CpG-specific, split by health status, VNTR genotype and season

| Female | Season |  |  |  |  |  | Health Status |  |  |  |  |  | uVNTR |  |  |  |  |  |
| --- | --- | --- | --- | --- | --- | --- | --- | --- | --- | --- | --- | --- | --- | --- | --- | --- | --- | --- |
|  | Spring / Summer |  |  | Autumn / Winter |  |  | PAT |  |  | HC |  |  | High |  |  | Low |  |  |
|  | N | mean | SD | N | mean | SD | N | mean | SD | N | mean | SD | N | mean | SD | N | mean | SD |
| Avg. | 24 | 0.503 | 0.090 | 20 | 0.441 | 0.069 | 18 | 0.471 | 0.073 | 26 | 0.477 | 0.095 | 22 | 0.473 | 0.066 | 22 | 0.476 | 0.104 |
| CpG1 | 19 | 0.585 | 0.221 | 18 | 0.576 | 0.254 | 15 | 0.707 | 0.227 | 22 | 0.495 | 0.202 | 18 | 0.527 | 0.181 | 19 | 0.632 | 0.270 |
| CpG2 | 24 | 0.346 | 0.107 | 20 | 0.243 | 0.084 | 18 | 0.274 | 0.096 | 26 | 0.317 | 0.116 | 22 | 0.305 | 0.093 | 22 | 0.294 | 0.125 |
| CpG3 | 23 | 0.496 | 0.158 | 20 | 0.389 | 0.119 | 17 | 0.373 | 0.131 | 26 | 0.493 | 0.145 | 22 | 0.441 | 0.152 | 21 | 0.451 | 0.152 |
| CpG4 | 23 | 0.473 | 0.115 | 20 | 0.424 | 0.104 | 17 | 0.467 | 0.106 | 26 | 0.440 | 0.115 | 22 | 0.459 | 0.101 | 21 | 0.441 | 0.123 |
| CpG5 | 24 | 0.249 | 0.115 | 20 | 0.181 | 0.065 | 18 | 0.205 | 0.088 | 26 | 0.228 | 0.109 | 22 | 0.218 | 0.080 | 22 | 0.219 | 0.120 |
| CpG6 | 24 | 0.474 | 0.130 | 20 | 0.400 | 0.118 | 18 | 0.438 | 0.124 | 26 | 0.442 | 0.135 | 22 | 0.441 | 0.120 | 22 | 0.440 | 0.140 |
| CpG7 | 24 | 0.570 | 0.143 | 20 | 0.473 | 0.115 | 18 | 0.506 | 0.122 | 26 | 0.540 | 0.149 | 22 | 0.537 | 0.121 | 22 | 0.516 | 0.156 |
| CpG8 | 24 | 0.437 | 0.148 | 20 | 0.375 | 0.110 | 18 | 0.389 | 0.125 | 26 | 0.423 | 0.141 | 22 | 0.397 | 0.085 | 22 | 0.421 | 0.172 |
| CpG9 | 24 | 0.621 | 0.114 | 20 | 0.521 | 0.114 | 18 | 0.561 | 0.112 | 26 | 0.585 | 0.132 | 22 | 0.589 | 0.080 | 22 | 0.561 | 0.156 |
| CpG10 | 24 | 0.584 | 0.110 | 20 | 0.537 | 0.111 | 18 | 0.571 | 0.096 | 26 | 0.557 | 0.124 | 22 | 0.578 | 0.069 | 22 | 0.547 | 0.143 |
| CpG11 | 24 | 0.179 | 0.086 | 20 | 0.169 | 0.082 | 18 | 0.163 | 0.085 | 26 | 0.182 | 0.083 | 22 | 0.152 | 0.067 | 22 | 0.196 | 0.094 |
| CpG12 | 24 | 0.994 | 0.023 | 20 | 0.987 | 0.041 | 18 | 0.991 | 0.035 | 26 | 0.991 | 0.031 | 22 | 0.987 | 0.039 | 22 | 0.995 | 0.025 |
| CpG13 | 24 | 0.530 | 0.160 | 20 | 0.470 | 0.180 | 18 | 0.494 | 0.168 | 26 | 0.508 | 0.174 | 22 | 0.528 | 0.143 | 22 | 0.477 | 0.193 |

**Table 2:** Mean *MAOA* promoter / exon I / intron I region DNA methylation levels in males (%), average and CpG-specific, split by health status, VNTR genotype and season.

| Male | Season |  |  |  |  |  | Health Status |  |  |  |  |  | uVNTR |  |  |  |  |  |
| --- | --- | --- | --- | --- | --- | --- | --- | --- | --- | --- | --- | --- | --- | --- | --- | --- | --- | --- |
|  | Spring / Summer |  |  | Autumn / Winter |  |  | PAT |  |  | HC |  |  | High |  |  | Low |  |  |
|  | N | mean | SD | N | mean | SD | N | mean | SD | N | mean | SD | N | mean | SD | N | mean | SD |
| Avg. | 7 | 0.161 | 0.089 | 22 | 0.122 | 0.026 | 11 | 0.153 | 0.071 | 18 | 0.118 | 0.025 | 22 | 0.133 | 0.057 | 7 | 0.126 | 0.016 |
| CpG1 | 7 | 0.039 | 0.102 | 19 | 0.000 | 0.000 | 9 | 0.030 | 0.090 | 17 | 0.000 | 0.000 | 20 | 0.014 | 0.060 | 6 | 0.000 | 0.000 |
| CpG2 | 7 | 0.024 | 0.064 | 22 | 0.003 | 0.012 | 11 | 0.017 | 0.051 | 18 | 0.003 | 0.013 | 22 | 0.011 | 0.037 | 7 | 0.000 | 0.000 |
| CpG3 | 7 | 0.218 | 0.114 | 21 | 0.204 | 0.181 | 11 | 0.250 | 0.218 | 17 | 0.180 | 0.119 | 21 | 0.184 | 0.139 | 7 | 0.278 | 0.224 |
| CpG4 | 7 | 0.051 | 0.136 | 22 | 0.002 | 0.010 | 11 | 0.037 | 0.108 | 18 | 0.000 | 0.000 | 22 | 0.016 | 0.077 | 7 | 0.006 | 0.017 |
| CpG5 | 7 | 0.032 | 0.057 | 22 | 0.020 | 0.059 | 11 | 0.046 | 0.085 | 18 | 0.009 | 0.027 | 22 | 0.028 | 0.066 | 7 | 0.009 | 0.017 |
| CpG6 | 7 | 0.059 | 0.155 | 22 | 0.004 | 0.019 | 11 | 0.045 | 0.124 | 18 | 0.000 | 0.000 | 22 | 0.023 | 0.089 | 7 | 0.000 | 0.000 |
| CpG7 | 7 | 0.071 | 0.150 | 22 | 0.019 | 0.045 | 11 | 0.060 | 0.125 | 18 | 0.014 | 0.034 | 22 | 0.037 | 0.093 | 7 | 0.016 | 0.037 |
| CpG8 | 7 | 0.053 | 0.101 | 22 | 0.001 | 0.005 | 11 | 0.026 | 0.080 | 18 | 0.006 | 0.025 | 22 | 0.018 | 0.060 | 7 | 0.000 | 0.000 |
| CpG9 | 7 | 0.096 | 0.166 | 22 | 0.028 | 0.059 | 11 | 0.086 | 0.139 | 18 | 0.019 | 0.048 | 22 | 0.058 | 0.109 | 7 | 0.003 | 0.004 |
| CpG10 | 7 | 0.075 | 0.155 | 22 | 0.006 | 0.023 | 11 | 0.040 | 0.125 | 18 | 0.012 | 0.035 | 22 | 0.030 | 0.092 | 7 | 0.001 | 0.002 |
| CpG11 | 7 | 0.045 | 0.059 | 22 | 0.012 | 0.027 | 11 | 0.022 | 0.037 | 18 | 0.019 | 0.041 | 22 | 0.022 | 0.042 | 7 | 0.014 | 0.028 |
| CpG12 | 7 | 0.945 | 0.086 | 22 | 0.912 | 0.089 | 11 | 0.937 | 0.070 | 18 | 0.910 | 0.098 | 22 | 0.913 | 0.098 | 7 | 0.944 | 0.040 |
| CpG13 | 7 | 0.381 | 0.074 | 22 | 0.360 | 0.136 | 11 | 0.372 | 0.120 | 18 | 0.361 | 0.128 | 22 | 0.370 | 0.128 | 7 | 0.351 | 0.114 |

**Figure 1:**
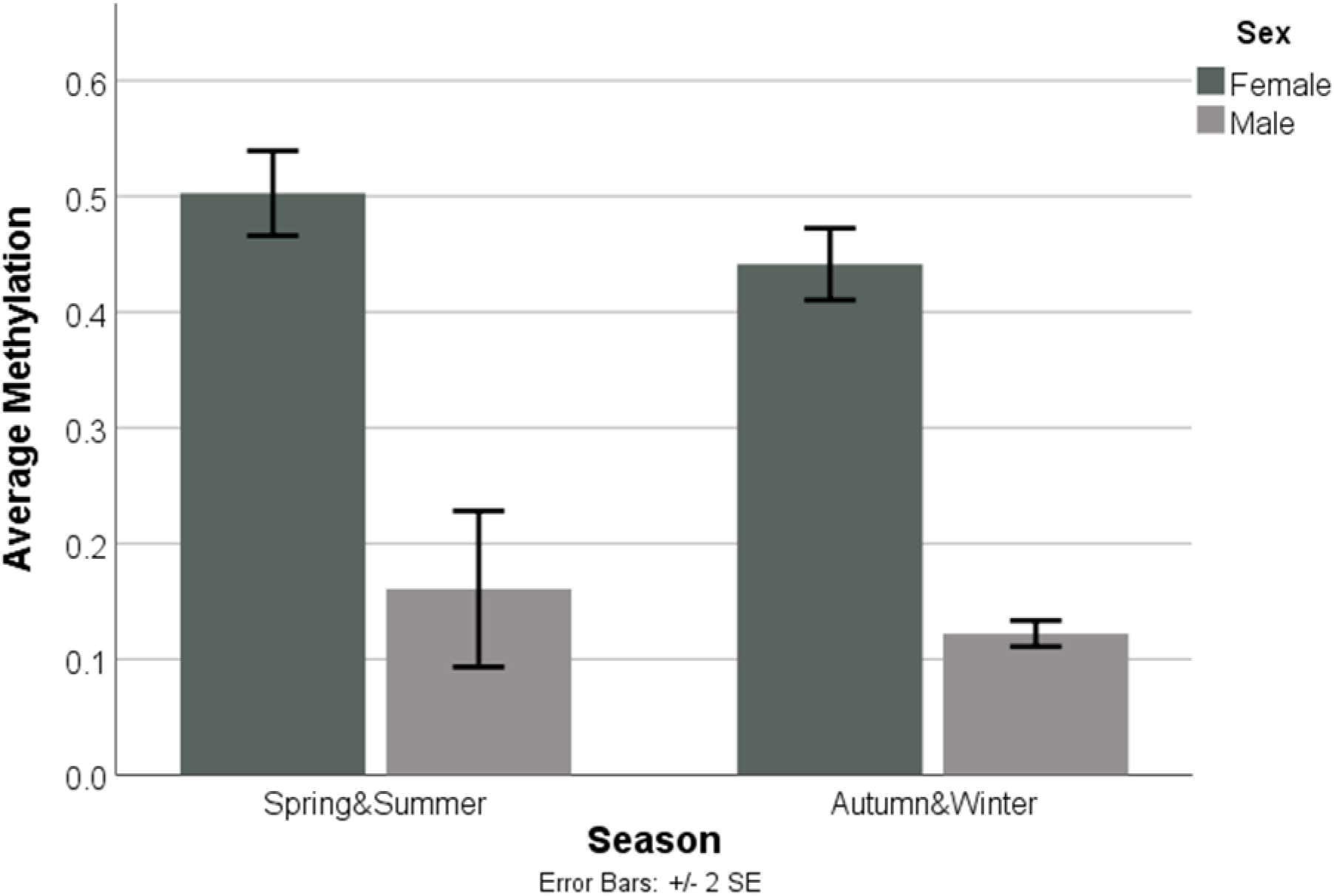
Lower average promoter / exon I / intron I region DNA methylation in autumn and winter compared with spring and summer within the female subject group (two-sided Mann-Whitney U-test, p_uncorr =_ 0.030).

### 3.2 Effect of *MAOA* DNA methylation on MAO-A V_T_

Initial GLM implemented to detect potential covariates (excluding methylation data) revealed no significant effect of health status, season or VNTR genotype on MAO-A V_T_. However, an effect of sex on MAO-A V_T_ could be found, with higher MAO-A V_T_ in females when compared to males (p_uncorr_ = 0.033).

Average *MAOA* DNA methylation had no significant effect on MAO-A V_T_, regardless of whether health, sex or season were corrected for via inclusion as covariates.

Furthermore, we investigated potential CpG site-specific effects of the abovementioned covariates on MAO-A V_T_ in an exploratory manner and found an effect of sex at CpG 3 (p_uncorr_ = 0.035), 5 (p_uncorr_ = 0.040), 6 (p_uncorr_ = 0.032), 8 (p_uncorr_ = 0.027) and 11 (p_uncorr_ = 0.011). Analyses on ROI-specific MAO-A VT revealed no effects of average *MAOA* DNA methylation.

## 4 Discussion

Here, we explored the effect of *MAOA* DNA methylation on global brain MAO-A V_T_ in healthy controls and patients with SAD. In contrast to prior findings on the effect of *MAOA* promoter methylation on cerebral MAO-A levels (23), no statistically significant association was observed for the 13 CpG sites assessed here (promoter / exon I / intron I), nor their average methylation. In addition to well-known sex differences in methylation of X-chromosomal genes (37) such as *MAOA*, we detected a statistically significant effect (uncorrected) of season on *MAOA* DNA methylation with higher methylation levels in spring / summer. Health status (SAD patients vs. controls) and *MAOA* promoter VNTR genotype (grouped by high / low expressing alleles) did not affect average *MAOA* DNA methylation. However, exploratory analyses of CpG-specific methylation revealed effects of health status and season for some CpGs.

### 4.1 Analysis of *MAOA* DNA methylation

We observed higher *MAOA* methylation in samples that were collected during spring or summer in female subjects. Although this association did not survive correction for multiple comparisons, it is in accordance with the role of season within the serotonergic system (38), including MAO-A (28), and suggests increased serotonin availability during spring / summer as conferred by lower MAO-A activity due to *MAOA* hypermethylation. Restriction of this effect to the female sex is potentially based on the aforementioned X-linked nature of *MAOA* and general consequences for methylation (37, 39). As reviewed by Nugent and McCarthy, DNA methylation, especially of X-chromosomal genes, follows a sex-specific pattern (37). This can be traced back to the transcriptional inactivation of one X-chromosome in mammalian female cells (46, XX), also referred to as X silencing (39) in order to balance the excess of X-linked alleles (40, 41). This results in higher average methylation levels of, e.g., *MAOA* in females (42, 43).

In general, epigenetic adaptations such as DNA methylation are known to be sensitive to a variety of environmental influences (44) ranging from drugs (45), prenatal or chronic adult exposure to environmental stimuli (46) such as air pollution (47) and toxins (48–50) or tobacco smoke (51, 52), as well as to nutritional factors (53). Furthermore, Bind and colleagues observed that environmental temperature and relative humidity were associated with dynamic changes in DNA methylation (54). In line with this assumption, Ricceri et al. reported higher mean methylation in spring and summer for certain genes (LINE-1, *RASSF1A* and *MGMT*) when compared to samples collected in autumn or winter (55). Notably, such findings are not limited to humans, with changes to methylation observed in hibernating animals (56, 57). Thus, factors that follow a seasonal pattern (e.g., outdoor temperature, exposure to air pollutants and ozone (58), seasonal changes to the circadian rhythm (59)), likely affect methylation. In addition, seasonal changes to nutrition such as folate and flavonoid intake may also play a role (60, 61). Expression and function of various proteins within the serotonergic system were shown to be sensitive to season and light (26, 62–65). Based on our methylation results, one might hypothesize that epigenetic processes facilitate these seasonal protein-level changes. However, seasonal effects on methylation did not carry over to changes in MAO-A V_T_ expression in our study, suggesting that these effects are minor when compared to other environmental variables (66) or pathologic conditions known to affect MAO-V_T_ (35).

Average methylation was utilized as a primary outcome parameter in accordance with previous clinical studies. However, based on middle to low inter-correlation between methylation of specific, particularly outer, CpGs (see supplement), CpG site specific analysis was performed (42). These exploratory analyses demonstrated an effect of health status (patients vs. controls) on methylation levels of individual *MAOA* CpG sites in each sex as well as an effect of season in females. Thus, these findings are partially in accordance with the effect of season on average methylation and point towards potential changes within SAD. However, despite prior findings on the pathophysiological relevance of the pre-defined *MAOA* CpG sites in our sample (16, 20), the number of tests CpG-specific assessments require necessitates investigations in a larger sample.

Again, we did not detect evidence for a significant effect of SAD on average *MAOA* DNA methylation. As summarized by Domschke and Ziegler (17), changes to average *MAOA* methylation of the genetic subregion we assessed were shown in psychiatric conditions (24, 67). Furthermore, Peng et al. investigated changes in *MAOA* promoter region methylation in depressive states, showing that methylation was negatively associated with depressive symptoms (12). Thus, in conjunction with this literature, our study is suggestive of a different or lesser role of *MAOA* methylation in SAD pathophysiology than it is the case for non-seasonal depression. On a theoretical level, this is in accordance with authors that promote the concept of SAD as an individual entity, rather than a subtype of major depressive disorder (68).

Finally, we did not find an effect of *MAOA* VNTR genotype on average *MAOA* DNA methylation of the amplicon comprising promoter / exon I / intron I. VNTR was taken into consideration based on its pronounced effect on MAO-A function (31) and thus potential impact on MAO-A V_T_, methylation, or both. An association of VNTR genotype and *MAOA* DNA methylation levels has been discussed (69), thought this is contradicted by others (23). However, it should be taken into account that compared to the presently investigated promoter / exon I / intron I region, the VNTR sequence is located further 5’ within the promoter of *MAOA*. Thus, any effects would be indirect, for example via secondary effects on other regulatory processes that include the promoter.

### 4.2 Effect of *MAOA* DNA methylation on MAO-A V_T_

One previous study assessed the effect of peripheral *MAOA* methylation (23) on brain MAO-A levels using [^11^C]clorgyline (23). The authors reported an association between CpG site specific *MAOA* core promoter methylation further 5’ and brain MAO-A levels in healthy individuals. Here, we assessed methylation within a promoter / exon I / intron I region, based on previous literature demonstrating altered methylation of this sequence in psychiatric disorders (16, 20) and prior observations that exon I methylation in general may result in particularly strong downregulation of transcription (21). As increased MAO-A is understood as an endophenotype of affective disorders (4), we postulated that altered DNA methylation may facilitate changes in MAO-A V_T_ previously observed in depression. The lack of an association in our study may thus result from the methylation sites we assessed and could be suggestive of a stronger association between promoter methylation and cerebral MAO-A. However, the choice of tracer may also underlie the observed differences in findings between studies. These involve differences between [^11^C]clorgyline and [^11^C]harmine in enzyme binding specifity as well as kinetics, with [^11^C]harmine showing markedly higher MAO-A specifity (70). Moreover, in contrast to [^11^C]harmine, [^11^C]clorgyline binds irreversibly to MAO (71) and exhibits characteristics that may limit data quality, including the presence of radioactive metabolites with MAO affinity (72). Importantly, [^11^C]harmine has developed as the most commonly utilized radioligand for brain MAO-A imaging in psychiatry (4, 29, 35, 73–75). Therefore, further investigations with a comparable study design are needed to elucidate the role of *MAOA* DNA region in the relationship between methylation and cerebral MAO-A. Moreover, a wide range of post-transcriptional processes regulate protein levels of serotonergic proteins, including MAO-A, as illustrated by recent studies demonstrating only weak or no association between mRNA and protein levels (75–77). Importantly, Komorowski et al. found a significant link between gene and protein expression for certain serotonin receptors (5-HT_1A_ and 5-HT_2A_). In contrast, only a weak association was shown for MAO-A when PET binding values and gene as well as protein expression were correlated in a voxel- and region-wise analysis (76). These factors, which cannot be addressed within our study design, may obscure the effects of methylation and other epigenetic processes on cerebral MAO-A V_T_.

### 4.3 Limitations

The following limitations should be addressed. Analysis of *MAOA* DNA methylation requires a sex-specific approach, which results in smaller sample sizes. As methylation is susceptible to a wide variety of influences (44, 46), variation is high, potentially obscuring smaller effect sizes. As a result, many studies of x-linked genes limit testing to male subjects (23). However, assessment in both sexes is nevertheless important for a more comprehensive understanding of x-linked genetic and epigenetic processes. The study at hand assesses methylation of a DNA region (*MAOA* promoter / exon I / intron I) located further 3’ than it was the case in the study performed by Shumay et al. (23). This decision was hypothesis-driven, based on the region’s clinical implications and potentially potent effect on protein expression (13, 16, 20, 78). However, it limits comparability to prior findings that focus on the *MAOA* promoter (23). In addition, in order to utilize PET data with temporal proximity to the time point of methylation analysis, either PET1 or PET3 data was used. Thereby, some SAD patients were remitted at the time of assessment and some subjects had received either BLT or placebo. Thus, for some individuals, sustained effects of BLT on *MAOA* DNA methylation or MAO-A expression cannot be ruled out completely.

## Data Availability

All data produced in the present study are available upon reasonable request to the authors.

## 5 Conclusion

Here we aimed to assess the effect of *MAOA* promoter / exon I / intron I region DNA methylation (16, 20) on cerebral MAO-A V_T_ assessed with [^11^C]harmine PET. We also probed the influence of various demographic and environmental characteristics on *MAOA* methylation including sex, depression (seasonal affective disorder) season and *MAOA* VNTR genotype. We did not find evidence for an effect of *MAOA* promoter / exon I / intron I region DNA methylation on brain MAO-A V_T_, in contrast to a previous PET study that demonstrated an association between *MAOA* promoter methylation and brain MAO-A levels. Thus, in comparison to a region within the promoter located further 5’, *MAOA* promoter / exon I / intron I region DNA methylation only appears to have limited impact on brain protein levels. Importantly, the use of different radiotracers needs to be taken into account. We observed an effect of season on average methylation in females, which is in accordance with extensive evidence for seasonal changes within the serotonergic system.

## 6 Acknowledgements

This research was funded by the Austrian Science Fund (FWF, KLI 504, PI: R. Lanzenberger; P24359, PI: D. Winkler). M. Murgaš is funded by the FWF (DOC 33-B27, Supervisor R. Lanzenberger). We want to express our thanks to M. Hienert, G. Gryglewski, T. Vanicek, A. Komorowski, A. Kautzky, L. Silberbauer J. Unterholzner, M.G. Godbersen, C. Kraus and S. Kasper for clinical assistance, A. Hahn, G.M. James, S. Ganger and M. Kloebl for technical support, G. Kranz, B. Spurny, S. Avramidis and J. Jungwirth for administrative help as well as J. Peters and K. Einenkel for data acquisition. We would also like to thank N. Berroterán-Infante, T. Balber, C. Philippe and M. Mitterhauser for assistance with radioligand synthesis, K. Rebhan for radioligand metabolite processing, T. Zenz and A. Krcal for PET technical assistance, and I. Leitinger, H. Ibeschitz, V. Weiss, and V. Pichler for assistance with PET measurements.

