## Supplemental Figure 1-4 for "Effect of *MAOA* DNA methylation on human *in vivo* protein expression measured by [^11^C]harmine PET in healthy and depressed individuals"

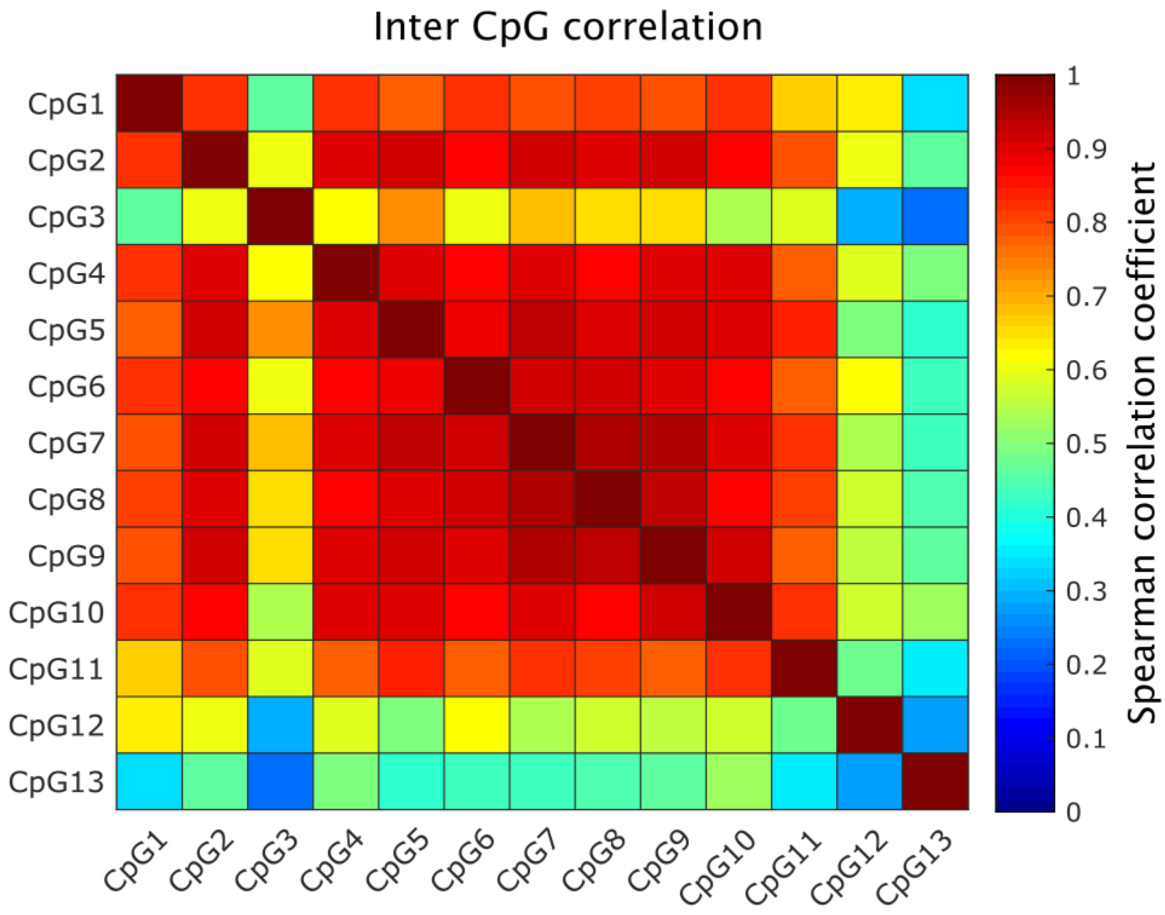


Figure S1: Correlation of mean methylation across all subjects between individual CpGs. Color chart represents spearman correlation coefficients.

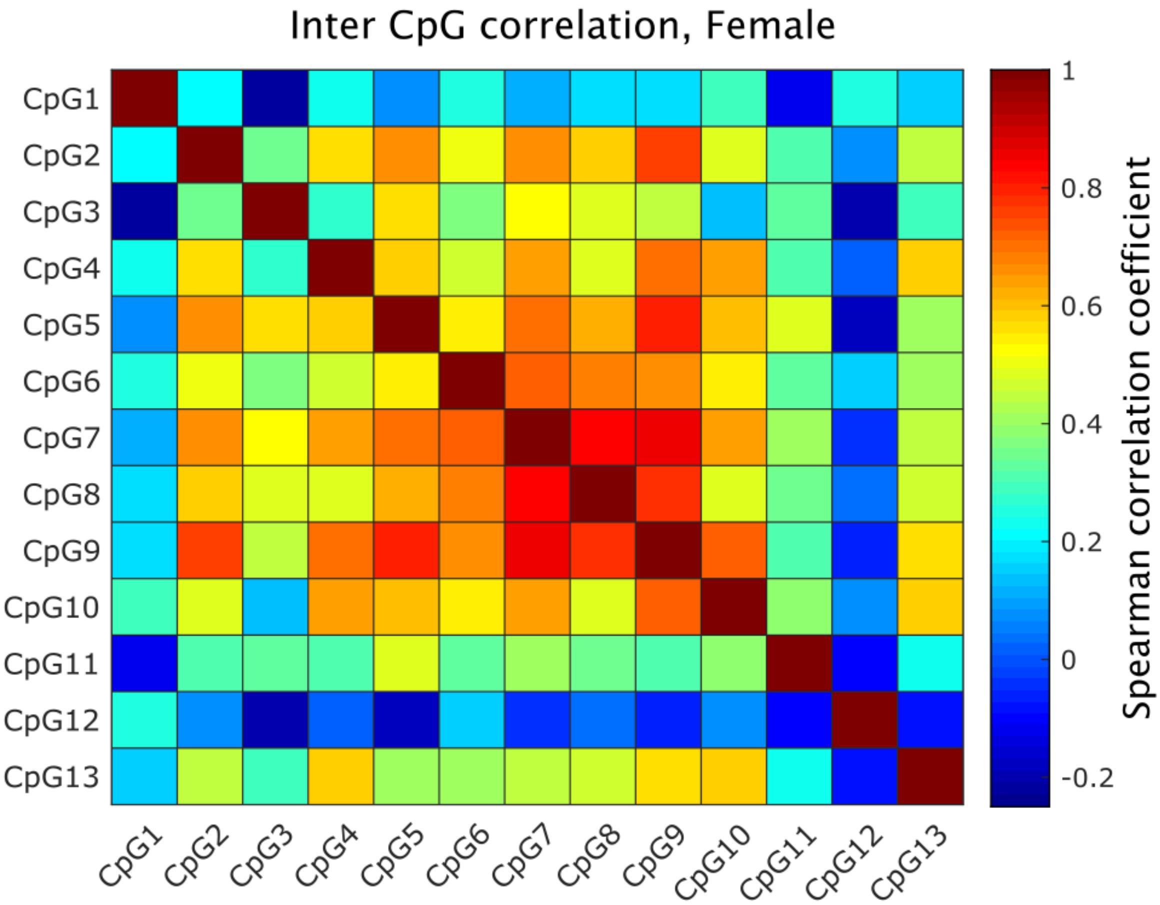


Figure S2: Correlation of mean methylation across female subjects between individual CpGs. Color chart represents spearman correlation coefficients.

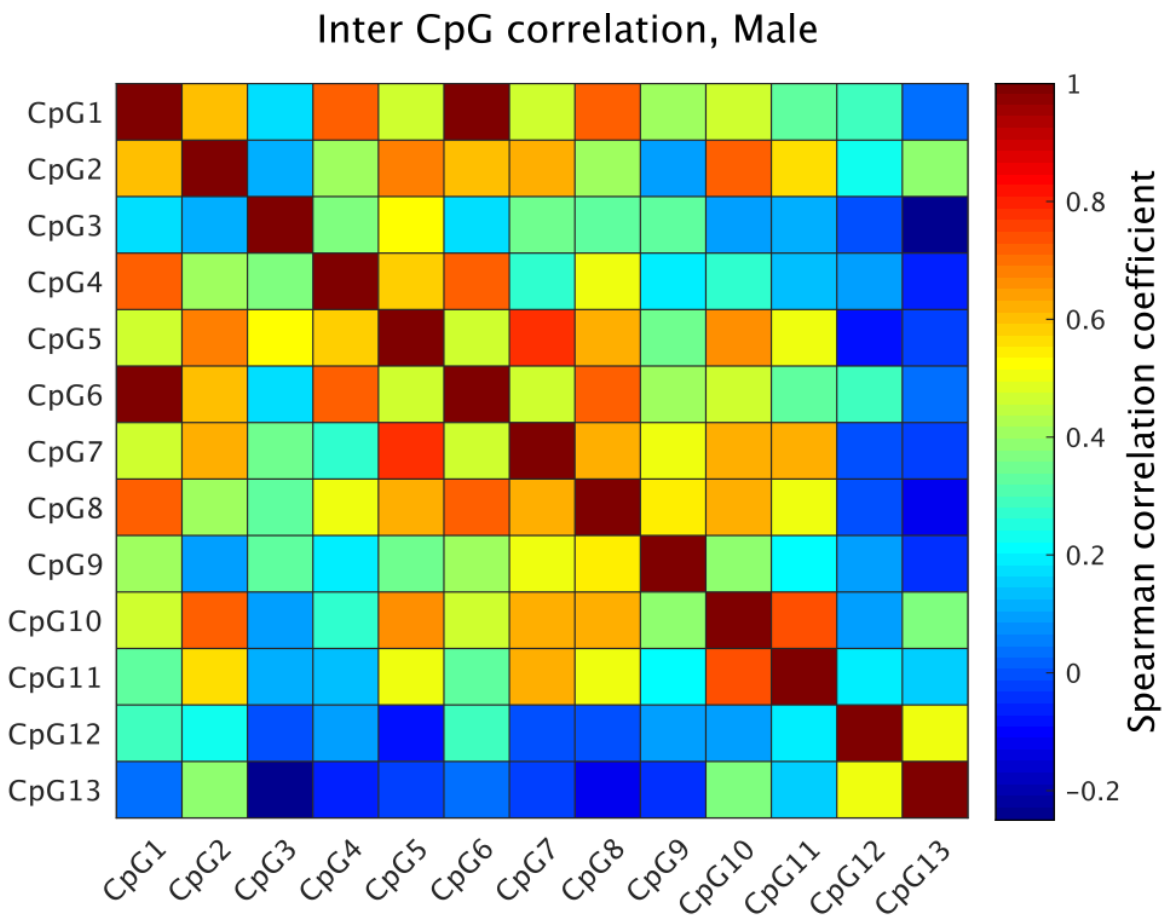


Figure S3: Correlation of mean methylation across male subjects between individual CpGs. Color chart represents spearman correlation coefficients.

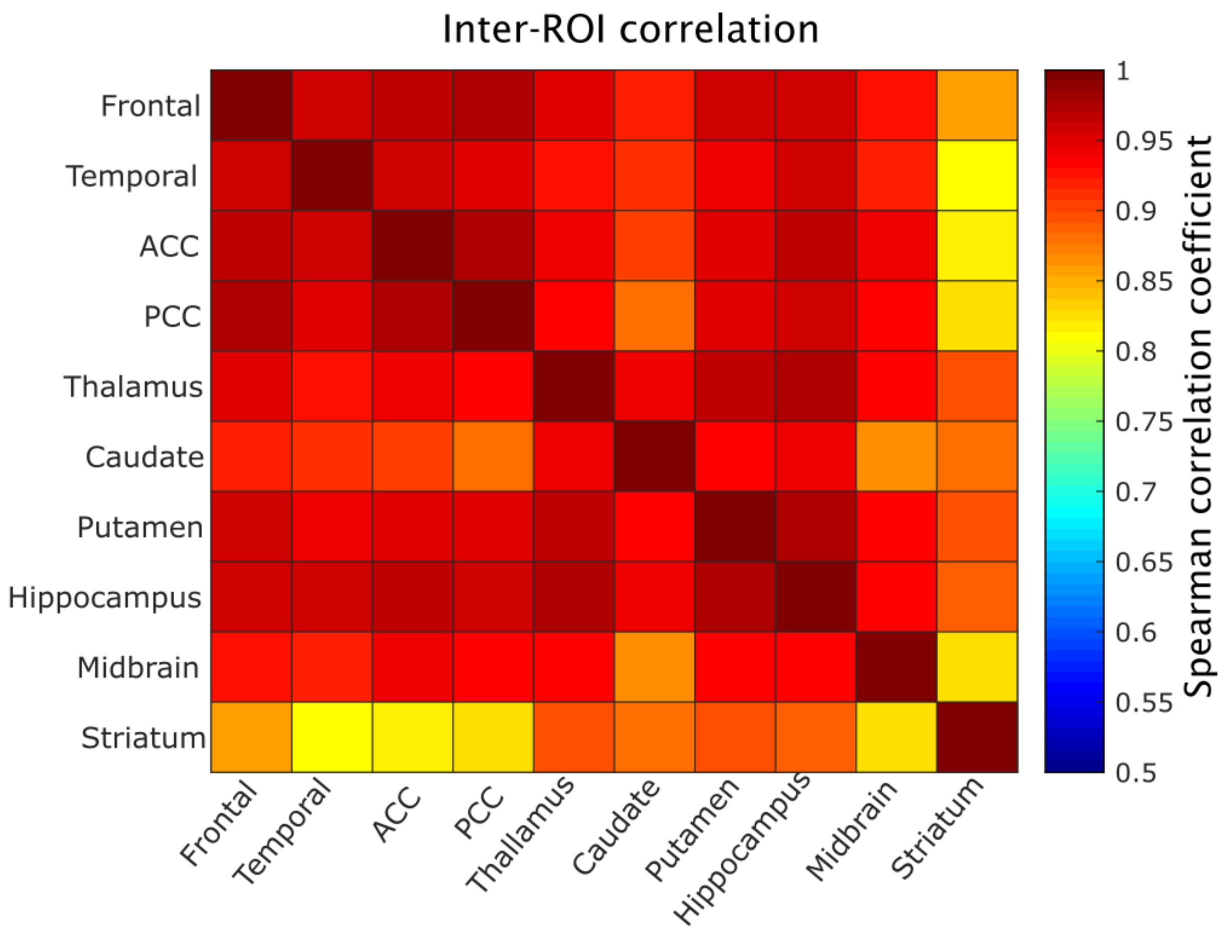


Figure S4: Correlation of mean MAO-A $V_T$ across subjects between individual ROIs. Color chart represents spearman correlation coefficients.
